# In Vivo Test-retest Quantitative Characterization of Echo Planar Imaging Apparent Diffusion Coefficient Reproducibility for Head and Neck Cancers on a 1.5T MR-Linac Platform: Technical Validation using QIBA Metrology

**DOI:** 10.1101/2025.03.06.25323426

**Authors:** Brigid A. McDonald, Dina El-Habashy, Renjie He, Sam Mulder, Sarah Mirbahaeddin, Abdallah S. R. Mohamed, Sara Ahmed, Yao Ding, Jihong Wang, Stephen Y. Lai, Alex Dresner, John Christodouleas, Clifton D. Fuller

## Abstract

**Background and Purpose:** To detect changes in apparent diffusion coefficient (ADC) values during radiation therapy for biological image-guided adaptive radiation therapy, the variability in ADC must be characterized. We evaluated the reproducibility of ADC values in head and neck cancers on a 1.5T MR-linac.

**Methods:** 39 head and neck cancer patients (36 primary tumors, 55 lymph nodes) were imaged with echo-planar imaging diffusion-weighted MRI on a 1.5T MR-linac at two time points before the start of radiation therapy. Mean and median ADC values and volume were measured for each lesion. Absolute and percent reproducibility coefficients (RC) were calculated. Linear regression analyses and F-tests were performed to determine whether lesion volume or time between scans impacted reproducibility.

**Results:** For primary tumors & lymph nodes: mean ADC, median ADC, and volume were 1.27 ± 0.33 mm^2^/s & 1.17 ± 0.34 mm^2^/s, 1.25 ± 0.35 & 1.16 ± 0.37 mm^2^/s, and 8.8 ± 12.3 cm^3^ & 6.5 ± 7.2 cm^3^, respectively. RC values of mean ADC were 0.355 mm^2^/s & 0.355 mm^2^/s for tumors & nodes, and %RC values were 29.1% & 31.1%; values were very similar for median ADC. Reproducibility was not significantly correlated with either volume or scan interval, but a trend of poorer reproducibility in smaller volumes was observed.

**Conclusion:** Considering previous reports that the optimal %ΔADC threshold for response prediction in head and neck cancers is around 15-30%, this sequence on the MR-linac has acceptable reproducibility for detecting larger ADC changes but may still miss some clinically significant changes.

## Introduction

Head and neck cancers, which account for approximately 4.7% of all cancers globally, affect more than 946,000 individuals each year [1]. These malignancies are commonly treated with radiation therapy, but patient outcomes vary widely [1–4], and many patients experience severe side effects such as xerostomia, dysphagia, and mucositis [5–7]. This variability underscores the need for personalized treatment approaches, which could potentially enhance tumor response while minimizing side effects for individual patients. Quantitative imaging, particularly diffusion-weighted MRI (DWI), has emerged as a promising tool for predicting radiation therapy outcomes [8,9]. Early changes in the apparent diffusion coefficient (ADC) during treatment are especially promising as predictors of treatment response [10–18].

The advent of hybrid MR-linac systems has revolutionized radiation therapy by enabling daily adaptive on-line treatment [19]. The Elekta Unity system not only allows precise radiation delivery but also integrates a 1.5T MRI scanner, offering the capability to obtain serial DWI scans throughout the treatment course. These scans can assess treatment response, potentially guiding personalized therapy [20,21]. However, this approach hinges on the use of robust DWI sequences to accurately capture changes in tumor diffusion as a surrogate of treatment efficacy. The Unity MR-linac system differs from conventional MRI scanners in several key ways that impact DWI acquisitions: 1) the split gradient coil design for radiation beam passage may lead to gradient non-linearities; 2) the lower gradient strength and slew rate can reduce signal-to-noise ratio by necessitating longer diffusion times for equivalent b-values; and 3) the relatively low number of channels (2×4) in the system’s body coil further reduces signal-to-noise ratio [22–25]. Despite these differences, studies have shown that properly optimized DWI sequences on the MR-linac can perform similarly to clinical sequences on conventional 1.5T MRI scanners [26,27].

To ensure the reliability of DWI as a quantitative imaging biomarker, the Quantitative Imaging Biomarker Alliance (QIBA) recommends using test-retest studies to assess the in vivo repeatability and reproducibility of these measurements [28]. According to guidelines by Obuchowski and Bullen, a minimum sample size of 35 subjects is required to accurately quantify the repeatability and reproducibility coefficients of variation [29]. QIBA has published consensus ADC profiles for brain, liver, prostate, and breast cancers, which provide performance claims based on pooled analyses of multiple test-retest studies in each disease site [30]. In this study, we expand on these efforts to quantify the reproducibility of ADC measurements in head and neck cancer primary tumors (n=36) and lymph nodes (n=55) using an echo-planar imaging DWI sequence on a 1.5T MR-linac.

## Methods

### Patients and Informed Consent

This retrospective study included HNC patients with histologically confirmed HNC who were imaged with echo planar imaging (EPI)-DWI on the 1.5T MR-linac (Elekta Unity; Stockholm, Sweden) at two or more time points between the pre-treatment MR simulation and first day of treatment (inclusive). If more than two eligible MRI examinations were available, the pair with the shortest time between scans was used. Six additional patients were included who had one pre-treatment DWI time point and the other DWI time point during the second (n=5) or third (n=1) fraction of a standard-fractionation treatment regimen (28-33 total fractions). A total of 47 patients were identified, and 8 were excluded for the following reasons: induction chemotherapy given between time points, previous radiation, primary tumor outside DWI field of view, and/or different b-values used. The final cohort consisted of 39 patients with a total of 36 primary tumors and 55 lymph nodes included in the analysis. Patient demographics are summarized in Table 1 and provided for individual patients in Supplement 1. The median (range) number of days between imaging time points was 11 (1-19). Ethics approval for this study was obtained through a retrospective data collection protocol with a waiver of informed consent (University of Texas MD Anderson Cancer Center Institutional Review Board protocol number RCR03-0800).

**Table 1.** Patient demographics.

| **Patient Characteristics** |  |
| --- | --- |
| **Age** |  |
| Median (range) | 64 (47-80) |
| **Gender** |  |
| Male | 36 |
| Female | 3 |
| **Primary Site** |  |
| Oropharynx (tonsil) | 14 |
| Oropharynx (base of tongue) | 18 |
| Larynx (glottis) | 3 |
| Nasopharynx | 1 |
| Parotid gland | 1 |
| Nasal cavity | 1 |
| Unknown primary | 1 |
| **Tumor Laterality** |  |
| Left | 17 |
| Right | 20 |
| Bilateral | 2 |
| **T Stage** |  |
| T1 | 17 |
| T2 | 13 |
| T3 | 5 |
| T4 | 3 |
| Tx (unknown primary) | 1 |
| **N Stage** |  |
| N0 | 11 |
| N1 | 21 |
| N2 | 5 |
| N3 | 2 |
| **M Stage** |  |
| M0 | 39 |
| **Overall Stage** |  |
| I | 25 |
| II | 8 |
| III | 2 |
| IVa | 3 |
| IVb | 1 |
| **HPV Status** |  |
| Positive | 32 |
| Negative | 1 |
| N/A | 6 |

### MRI Examinations

Patients were imaged in radiation therapy immobilization masks with a T2-weighted sequence and single-shot EPI-DWI (sequence parameters in Table 2) at two time points each. The majority of DWI images (n=63) had a 160 mm field of view in the z (slice) direction with repetition time (TR) = 5698 ms and a scan time of 4:51 (minutes: seconds). In some cases, the z direction field of view was manually changed, which in turn changed the TR and acquisition time (see Table 2). These differences in TR should have minimal impact on the ADC values, as ADC values have been shown to be stable across all TR values greater than 3000 ms [31,32]. ADC maps were automatically reconstructed on the scanner using only the b=150 and 500 s/mm^2^ images to minimize perfusion effects, per recommendations from the MR-Linac Consortium [22].

**Table 2.** MRI acquisition parameters. Abbreviations: TR = repetition time, TE = echo time, AP = anterior-posterior, RL = right-left, FH = foot-head.

|  | **T2-weighted** | **EPI-DWI** |
| --- | --- | --- |
| **Scan type** | 3D T2-weighted TSE | Single-shot SE EPI |
| **TR (ms)** | 1400 | 3613 (n=10)<br>5003 (n=4)<br>5698 (n=63)<br>7087 (n=1) |
| **TE (ms)** | 182 | 75 |
| **Flip angle (degrees)** | 90 | 90 |
| **Refocusing flip angle (degrees)** | 55 | N/A |
| **b-values (number of averages)** | N/A | 0 (2), 150 (4), 500 (12) |
| **Half scan factor** | N/A | 0.88 |
| **Fat suppression** | SPAIR | STIR |
| **Acceleration type (factor)** | SENSE (2) | SENSE (2.2) |
| **Phase encode direction** | AP | AP |
| **Field of view (RL x AP) (mm)** | 520 x 270 | 300 x 300 |
| **Field of view (FH) (mm)** | 200 | 100 (n=10)<br>140 (n=4)<br>160 (n=63)<br>200 (n=1) |
| **Acquisition pixel size (RL x AP x FH) (mm)** | 1.4 x 1.5 x 2 | 3.5 x 3.5 x 4 |
| **Reconstruction pixel size (RL x AP x FH) (mm)** | 0.81 x 0.81 x 2 | 1.56 x 1.56 x 4 |
| **Total scan duration (min:sec)** | 5:09 | 3:04 (n=10)<br>4:15 (n=4)<br>4:51 (n=63)<br>6:01 (n=1) |

### Image Segmentation and Data Analysis

Primary tumors and pathological lymph nodes were segmented by a radiation oncologist (D.M.E.) in VelocityAI (v3.0.1; Velocity Medical Solutions; Atlanta, GA) on the b0 images—which were rigidly registered to the T2w images as guidance for better visualization—then were rigidly copied to the ADC maps. Mean and median ADC values and volumes of each lesion were extracted using an in-house script. Wilcoxon signed rank tests (α=0.05) were performed in JMP (v15.0.0; SAS Institute Inc.; Cary, NC, USA) to determine if there were significant differences in volume, mean ADC, and median ADC between time points for PTs and LNs.

The following reproducibility metrics were calculated for both the mean ADC and median ADC according to QIBA consensus guideline procedures [28]: within-subject deviation (wSD), reproducibility coefficient (RC), within-subject coefficient of variation (wCV), and percent RC (%RC) where RC = 2.77*wSD and %RC = 2.77*wCV. In the QIBA formalism, the RC and %RC values are the threshold absolute change and percent change in ADC, respectively, that represent a real change in successive ADC measurements not attributed to the measurement error, with 95% confidence. The %RC should be used when the reproducibility depends on the magnitude of the ADC measurements. Briefly, wSD is calculated by

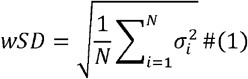

where N is the number of subjects (or lesions) and *σ*_*i*_ is the standard deviation of the two mean (or median) ADC measurements at the two time points for each subject. wCV is calculated by

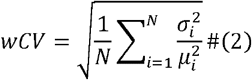

where *μ*_*i*_ is the mean of the two mean (or median) ADC measurements at the two time points for each subject.

Bland-Altman analysis was performed in GraphPad Prism (v10.0.3; GraphPad Software, LLC; Boston, MA) for PT and LN mean and median ADC values across time points. Plots were generated of the difference in ADC (time point 1 – time point 2) vs. average ADC between time points, and the mean bias and 95% limits of agreement were calculated. Next, we investigated whether the lesion volume and/or number of days between images impacted the reproducibility. Linear regression analyses and F-tests (α=0.05) were performed in JMP between these quantities and the wCV of each subject (or lesion). From equation (2), the wCV for each subject is calculated by

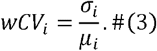

The average lesion volume between the two time points was used for the volume correlations.

## Results

Representative T2 and EPI-DWI images are shown in Figure 1. The volume, mean ADC, and median ADC of each lesion for each patient are given in Supplement 1. These values are summarized across the cohort in Table 3, and the reproducibility metrics for both mean and median ADC of the primary tumors and lymph nodes are included in Table 3 as well. The overall mean (± standard deviation) volume across both time points was 8.8 ± 12.3 cm^3^ for primary tumors and 6.5 ± 7.2 cm^3^ for lymph nodes. The overall mean (± standard deviation) of mean ADC across both time points was 1.27 ± 0.33 mm^2^/s for primary tumors and 1.17 ± 0.34 mm^2^/s for lymph nodes. The median ADC values were very similar to the mean ADC values: 1.25 ± 0.35 and 1.16 ± 0.37 mm^2^/s for primary tumors and lymph nodes, respectively. There were no statistically significant differences between time points for the volume or mean/median ADC values for primary tumors, but these values were significantly different for lymph nodes (p-values for all comparisons are in Table S1 (Supplement 2)). The RC and %RC values for mean ADC were 0.355 mm^2^/s and 29.1% for primary tumors and 0.355 mm^2^/s and 31.1% for lymph nodes, respectively. For median ADC, the RC and %RC values were 0.385 mm^2^/s and 31.9% for primary tumors and 0.366 mm^2^/s and 33.1% for lymph nodes, respectively.

**Table 3.**
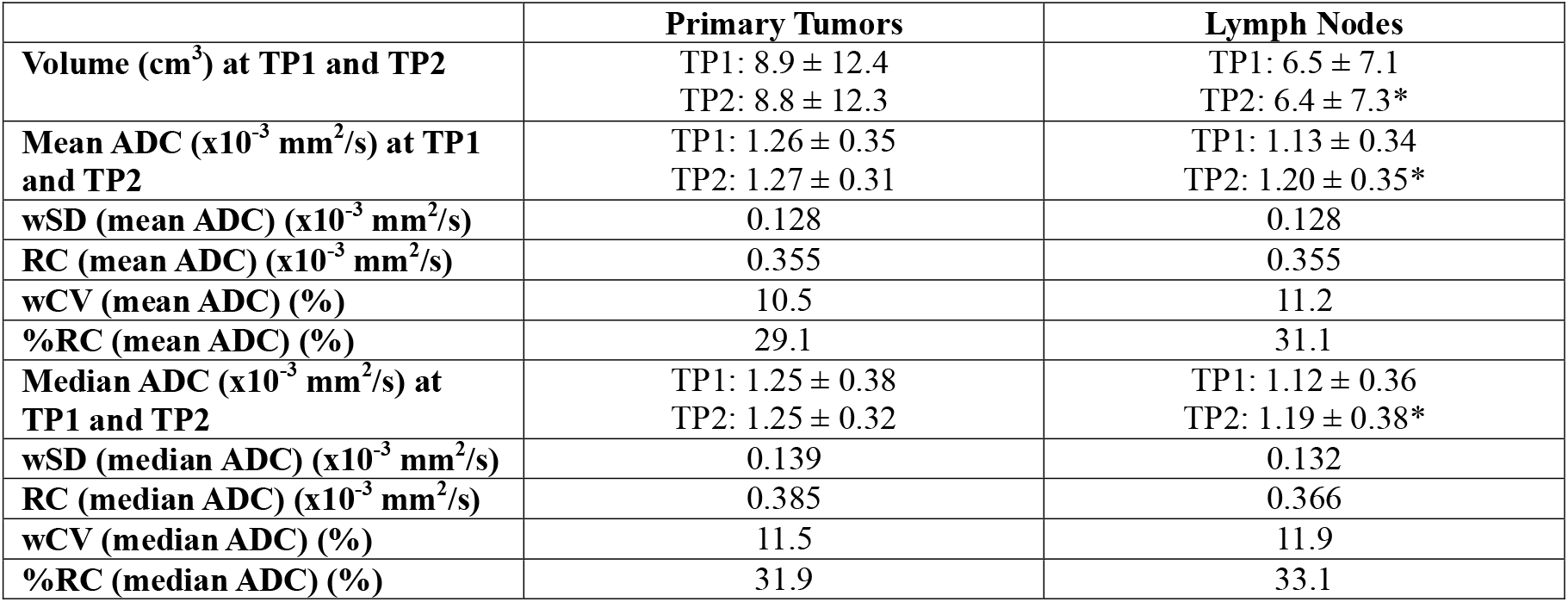
Volume, mean ADC, median ADC, and reproducibility metrics of both mean ADC and median ADC for primary tumors and lymph nodes. Volume and mean/median ADC values are expressed as the mean ± standard deviation over the cohort at each time point. Quantities that are statistically significantly different across time points are denoted with an asterisk after the second time point’s value. Abbreviations: TP = time point, ADC = apparent diffusion coefficient, wSD = within-subject deviation, RC = reproducibility coefficient, wCV = within-subject coefficient of variation, %RC = percent reproducibility coefficient.

|  | **Primary Tumors** | **Lymph Nodes** |
| --- | --- | --- |
| **Volume (cm<sup>3</sup>) at TP1 and TP2** | TP1: 8.9 ± 12.4<br>TP2: 8.8 ± 12.3 | TP1: 6.5 ± 7.1<br>TP2: 6.4 ± 7.3* |
| **Mean ADC (x10<sup>-3</sup> mm<sup>2</sup>/s) at TP1 and TP2** | TP1: 1.26 ± 0.35<br>TP2: 1.27 ± 0.31 | TP1: 1.13 ± 0.34<br>TP2: 1.20 ± 0.35* |
| **wSD (mean ADC) (x10<sup>-3</sup> mm<sup>2</sup>/s)** | 0.128 | 0.128 |
| **RC (mean ADC) (x10<sup>-3</sup> mm<sup>2</sup>/s)** | 0.355 | 0.355 |
| **wCV (mean ADC) (%)** | 10.5 | 11.2 |
| **%RC (mean ADC) (%)** | 29.1 | 31.1 |
| **Median ADC (x10<sup>-3</sup> mm<sup>2</sup>/s) at TP1 and TP2** | TP1: 1.25 ± 0.38<br>TP2: 1.25 ± 0.32 | TP1: 1.12 ± 0.36<br>TP2: 1.19 ± 0.38* |
| **wSD (median ADC) (x10<sup>-3</sup> mm<sup>2</sup>/s)** | 0.139 | 0.132 |
| **RC (median ADC) (x10<sup>-3</sup> mm<sup>2</sup>/s)** | 0.385 | 0.366 |
| **wCV (median ADC) (%)** | 11.5 | 11.9 |
| **%RC (median ADC) (%)** | 31.9 | 33.1 |

**Figure 1:**
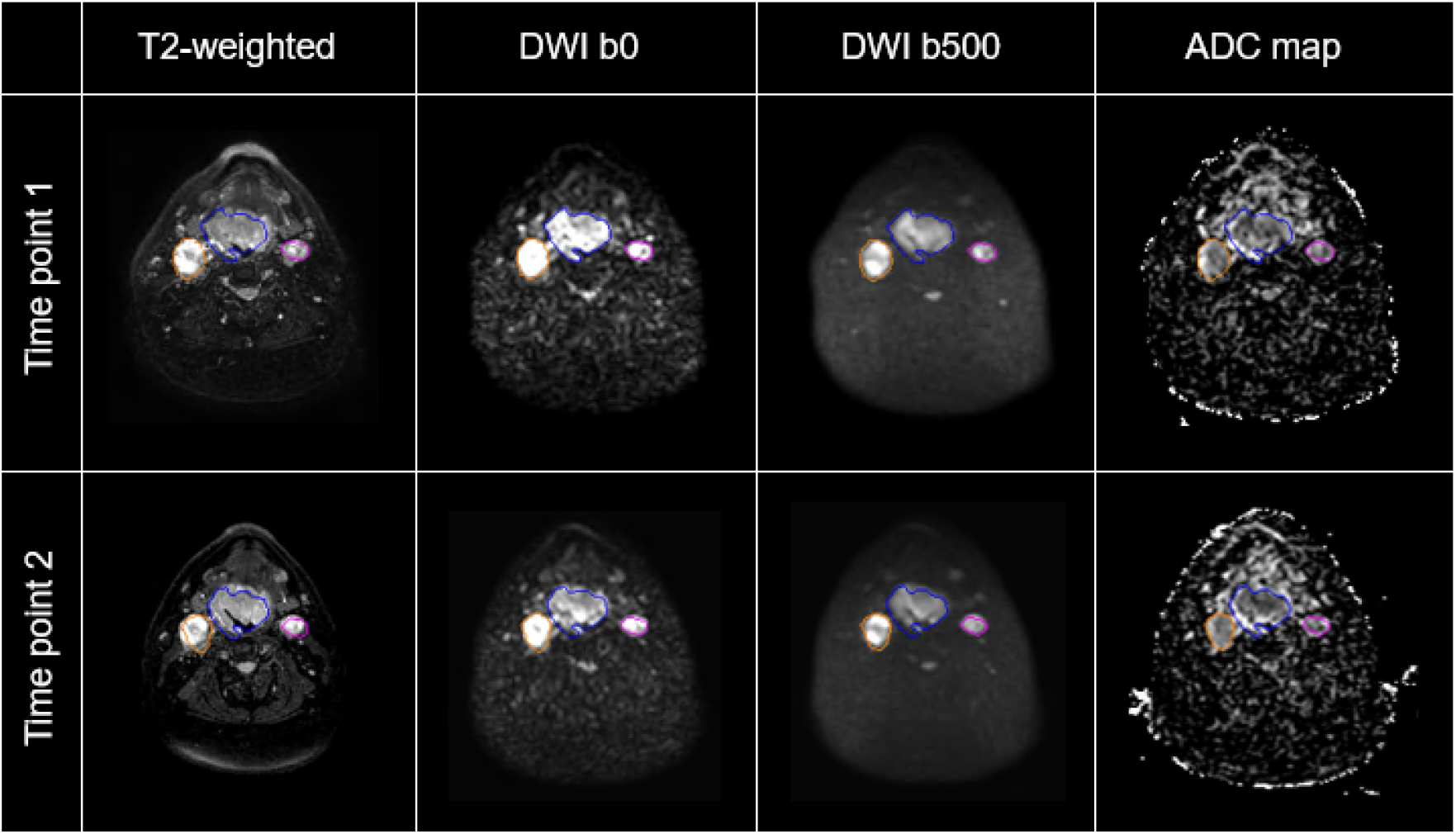
Representative T2 and EPI-DWI images (b = 0 and 500 s/mm^2^ images and ADC maps) for a patient at two time points. The primary tumor is segmented in blue, and two lymph nodes are segmented in orange and purple.

Bland-Altman plots are shown in Figure 2. The mean bias (time point 1 – time point 2) and limits of agreement for mean ADC were −0.00581 (−0.366 – 0.355) for primary tumors and −0.0645 (−0.400 – 0.271) for lymph nodes. For median ADC, mean bias and limits of agreement were −0.00101 (−0.392 – 0.390) for primary tumors and −0.0656 (−0.411 – 0.280) for lymph nodes.

**Figure 2:**
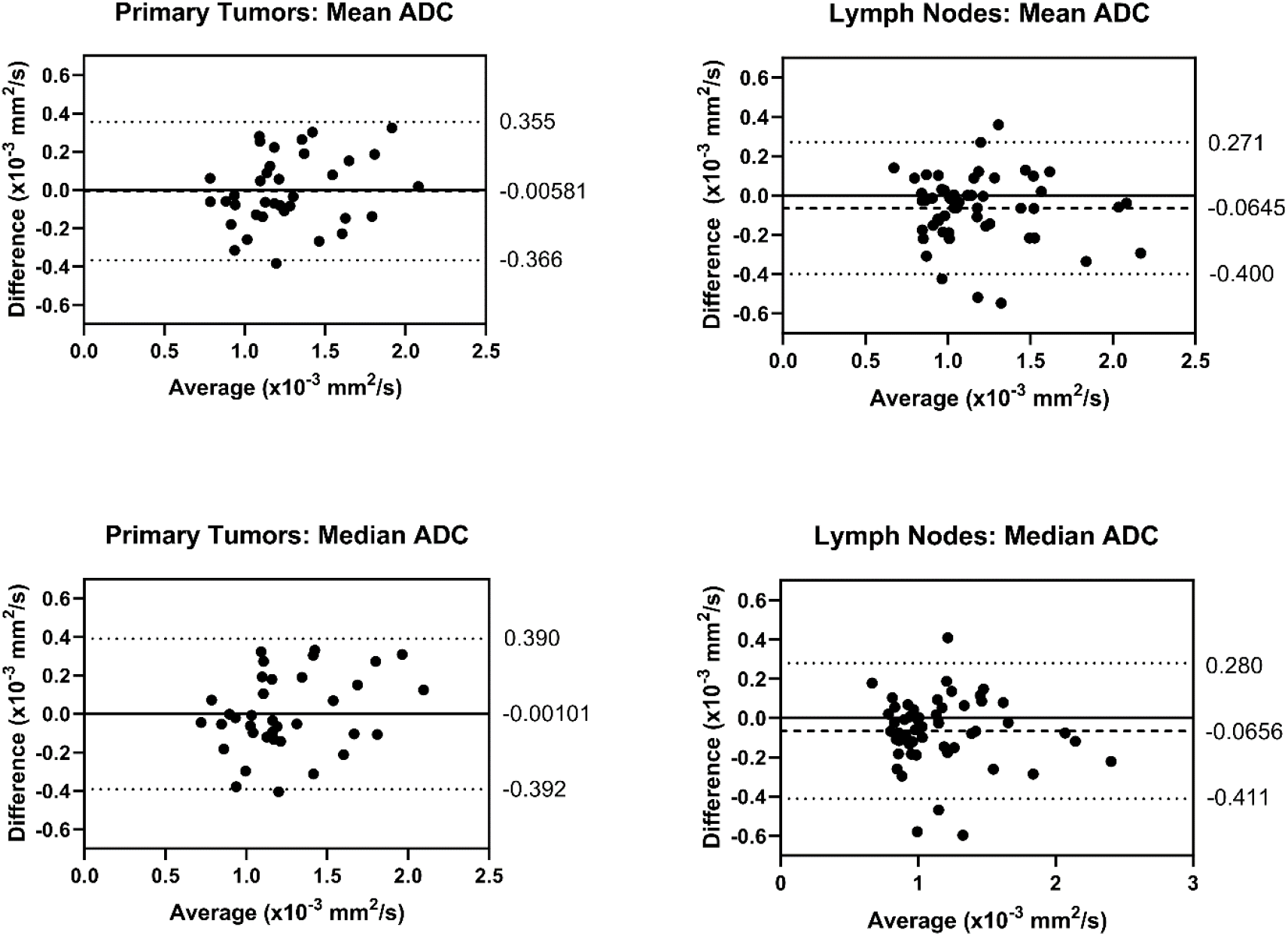
Bland-Altman plots representing the difference in ADC between time points (time point 1 – time point 2) vs. the average ADC between time points. Mean biases are represented by dashed lines, and limits of agreement are represented by dotted lines. The numerical values for the mean biases and limits of agreement are on the right side of each plot.

The results of the linear regression analyses (plots, r^2^ values, and p-values for all comparisons) are shown in Figures S1 and S2 (Supplement 2). Weak positive associations were found between wCV values (for both mean and median ADC) and days between scans for both primary tumors and lymph nodes. Weak negative associations were found between wCV values and lesion volumes for both primary tumors and lymph nodes. All comparisons showed weak associations between variables, with r^2^ values ranging from 0.0210 to 0.0655 across all comparisons. None of the F-tests for the slopes of these associations were statistically significant (i.e. none of the slopes were significantly non-zero). However, visual inspection of the plots for wCV vs. lesion volume (Figure S2) shows that most cases with high wCV values occurred for very small lesion volumes (below approximately 10 cm^3^).

## Discussion

In this study, we quantified the ADC reproducibility for head and neck cancers on a 1.5T MR-linac using the test-retest method [28]. This technical validation is a necessary pre-requisite step for biological image-guided adaptive radiation therapy, as it quantifies the measurement noise and sets the baseline threshold change in ADC that is detectable by this system and this DWI sequence. It has been well demonstrated that changes in mean ADC from baseline to week 2 or 3 of radiation therapy can predict various oncologic outcomes in head and neck cancers. From these studies, the reported optimal threshold change in ADC for outcome prediction typically ranges from about 15% to 30% [10–16] but has been reported as low as 7% [17] and as high as 53% [18] for different study cohorts. In this study, we calculated that the lowest detectable percent change in mean ADC with this EPI sequence on the 1.5T MR-linac is 29.1% for primary tumors and 31.1% for lymph nodes. This means that we may not be able to detect some clinically significant changes in ADC on this device without further optimization of the hardware and/or sequences to reduce noise and improve sensitivity. However, we can detect large ADC changes indicative of either very favorable or very poor response, which may still be informative for biological image-guided adaptive radiation therapy, depending on physician intent.

Few reports so far have quantified repeatability and/or reproducibility for head and neck cancers in either conventional MRI scanners [33,34] or MR-linacs [27,35], and the cohorts used in these studies were small (≤16 patients). With 36 primary tumors and 55 lymph nodes across 39 patients, our study has the largest cohort to date and is the first to reach the minimum recommended sample size of 35 patients for test-retest studies to determine the true repeatability/reproducibility coefficients [29].

Despite some key differences in methodology, the results of this study are similar to the two previous reports by Habrich et al. [35] and McDonald et al. [27], which both used EPI-DWI sequences with b-values of 150 and 500 s/mm^2^ on the 1.5T MR-linac. Habrich et al. acquired repeat DWI scans during multiple treatment fractions per patient (81 image pairs over 11 patients) with no repositioning between scans. For primary tumors and lymph nodes, they reported mean ADC repeatability coefficient values of 0.457 and 0.310 x10^−3^ mm^2^/s and percent repeatability coefficient values of 31.3% and 23.5%. In our group’s previous report [27], we had a smaller cohort (10 patients with two sets of repeat images per patient) but identical sequence parameters as the current study, and we measured both repeatability (same-day scans) and reproducibility (scans over multiple days) using mean ADC. We also reconstructed the ADC maps separately with b-values of 0 and 500 s/mm^2^ and with b-values of 150 and 500 s/mm^2^—the latter of which is recommended by the MR-Linac Consortium to minimize perfusion effects [22]—to determine whether the choice of b-values impacts the repeatability/reproducibility. We calculated percent reproducibility coefficients for primary tumors and lymph nodes of 52.5% and 39.5% for b=150,500 ADC maps and 28.5% and 27.6% for b=0,500 ADC maps. The values for b=150,500 ADC maps are larger than those reported in the current study (29.1% and 31.1% for primary tumors and lymph nodes) and by Habrich et al., but that study had the smallest sample size and thus may be more skewed by outliers.

We also investigated whether there was a difference in the reproducibility of mean ADC and median ADC values. In general, the median ADC should be less sensitive to outliers or voxel-level fitting errors than the mean ADC, and the median is typically preferred over the mean when distributions are non-Gaussian. Although the distributions of ADC values within head and neck lesions are generally non-Gaussian [15,36,37], the vast majority of studies looking at head and neck cancer ADC values used mean ADC and have found it to have predictive potential for tumor response [10–18]. In this study, the mean and median ADC values of the lesions were very similar; on average, the mean ADC was 0.02 x10^−3^ mm^2^/s higher than median ADC for primary tumors and 0.01 x10^−3^ mm^2^/s higher for lymph nodes. The RC and %RC values were slightly larger overall for median ADC than for mean ADC, but differences were small (0.03 x10^−3^ mm^2^/s for primary tumor RC, 0.011 x10^−3^ mm^2^/s for lymph node RC, and 2.0% for both primary tumor and lymph node %RC). These differences are clinically insignificant compared to the typical magnitude of ADC changes during radiation therapy, so the choice between mean and median ADC values should not significantly impact results.

One limitation of our study is that the scans were taken between 1-2 weeks apart for many patients (with a maximum of 19 days between scans) because they were imaged during routine MR simulation and MR-linac treatment appointments. The average increase in mean ADC between time points was 0.01 x10^−3^ mm^2^/s for primary tumors and 0.07 x10^−3^ mm^2^/s for lymph nodes, and the increase in ADC was statistically significant for lymph nodes in the Wilcoxon signed rank test. However, wCV values showed very weak positive associations with the days between repeat scans, with slopes that were not significantly different from zero in the F-test. Similarly, the limits of agreement from the Bland-Altman analysis were roughly symmetrical around zero, spanning from −0.400 x10^−3^ mm^2^/s to 0.271 x10^−3^ mm^2^/s for mean ADC of lymph nodes. These results suggest that the intervals between scans did not have a substantial impact on the reproducibility results in this cohort. We also investigated whether the lesion volume impacted the reproducibility. Although the negative association was also weak and non-significant, most outlier wCV values occurred for tumor or lymph node volumes below approximately 10 cm^3^, indicating that variability does increase for smaller lesions.

## Conclusion

In this study, we evaluated the reproducibility of ADC measurements in head and neck cancers using a 1.5T MR-linac with an echo-planar imaging DWI sequence. With the largest test-retest cohort to date for head and neck cancers, this study establishes an important baseline for future efforts in biological image-guided adaptive radiation therapy. Our findings show that, while the reproducibility of ADC for both primary tumors and lymph nodes is in line with previously reported values for the MR-linac, the system’s current limitations prevent the detection of smaller, clinically significant changes in ADC that could be used to predict treatment response early during the course of treatment. Further optimization of MR-linac hardware and sequences may enhance sensitivity, enabling more precise tracking of treatment response for biological image-guided adaptive therapy.

## Data Availability

All data produced in the present study are available upon reasonable request to the authors

https://doi.org/10.6084/m9.figshare.28548740.v1

## Declaration of Generative AI and AI-assisted technologies in the writing process

Statement: During the preparation of this work the authors used ChatGPT-4 (OpenAI; accessed in October 2024) to help write the Introduction section, specifically to turn detailed bulleted outlines into more eloquent text. All thoughts are original to the authors, and all references were added to this section without the use of ChatGPT. After using this tool, the authors reviewed and edited the content as needed and take full responsibility for the content of the publication.

