## Supplemental Material for "In Vivo Test-retest Quantitative Characterization of Echo Planar Imaging Apparent Diffusion Coefficient Reproducibility for Head and Neck Cancers on a 1.5T MR-Linac Platform: Technical Validation using QIBA Metrology"

**Supplemental Materials**

**Table S1:** Comparisons between time points for volume, mean ADC, and median ADC of primary tumors and lymph nodes. Values are expressed as mean ± standard deviation. Wilcoxon signed rank test p-values are shown in the last column (α=0.05). Abbreviations: TP = time point, ADC = apparent diffusion coefficient.

|  |  | **TP1 value** | **TP2 value** | **p-value** |
| --- | --- | --- | --- | --- |
| **Primary tumors** | **Volume (cm^3^)** | 8.9 ± 12.4 | 8.8 ± 12.3 | 0.0539 |
|  | **Mean ADC (x10^-3^ mm^2^/s)** | 1.26 ± 0.35 | 1.27 ± 0.31 | 0.7936 |
|  | **Median ADC (x10^-3^ mm^2^/s)** | 1.25 ± 0.38 | 1.25 ± 0.32 | 0.9387 |
| **Lymph nodes** | **Volume (cm^3^)** | 6.5 ± 7.1 | 6.4 ± 7.3 | 0.0432* |
|  | **Mean ADC (x10^-3^ mm^2^/s)** | 1.13 ± 0.34 | 1.20 ± 0.35 | 0.0060* |
|  | **Median ADC (x10^-3^ mm^2^/s)** | 1.12 ± 0.36 | 1.19 ± 0.38 | 0.0079* |

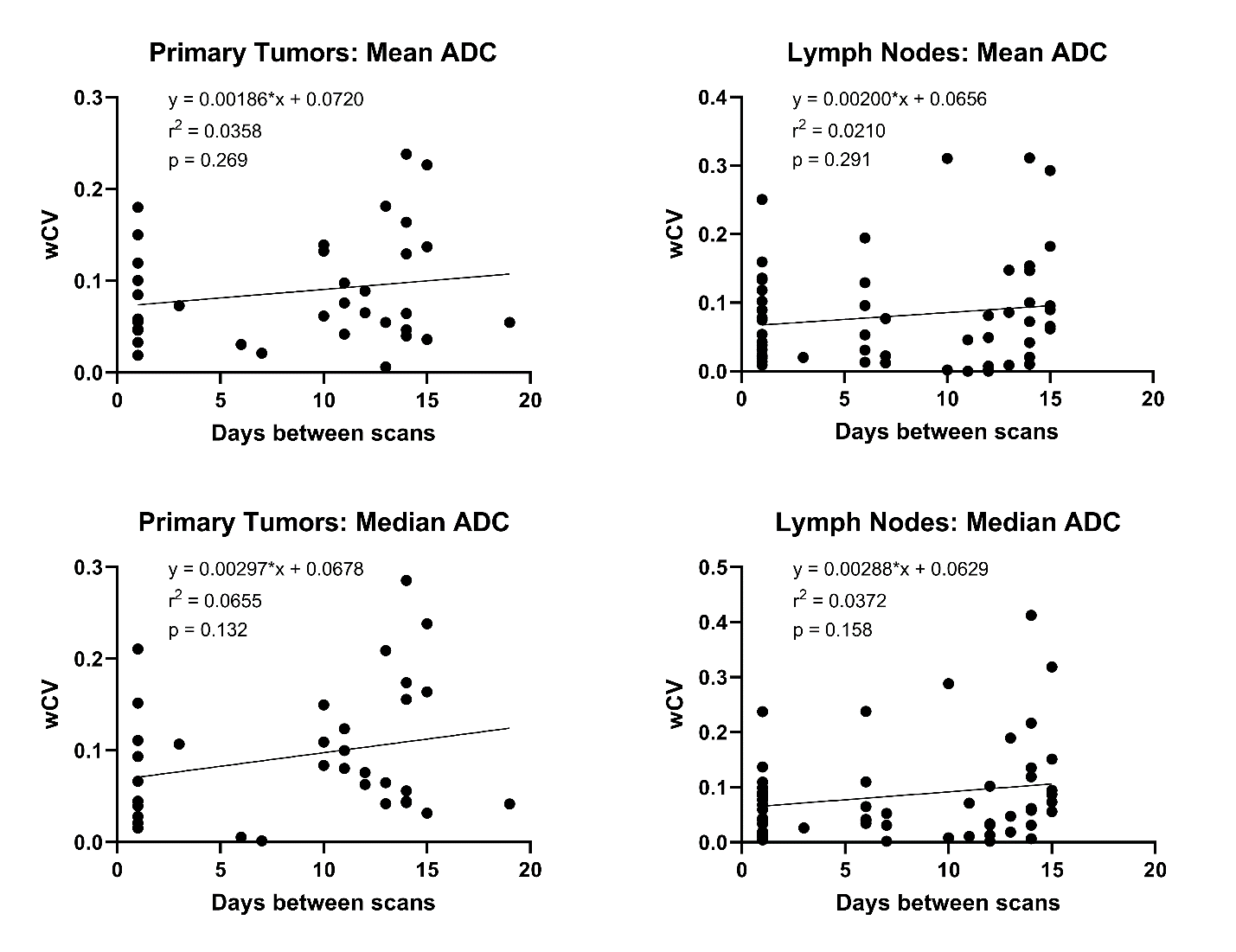

**Figure S1:** wCV vs. days between scans comparisons for both primary tumors and lymph nodes. wCV values using both mean ADC and median ADC were analyzed. The linear regression equation, r^2^ value, and p-value for the F-test (α=0.05) is shown for each comparison.

**Supplement 2**
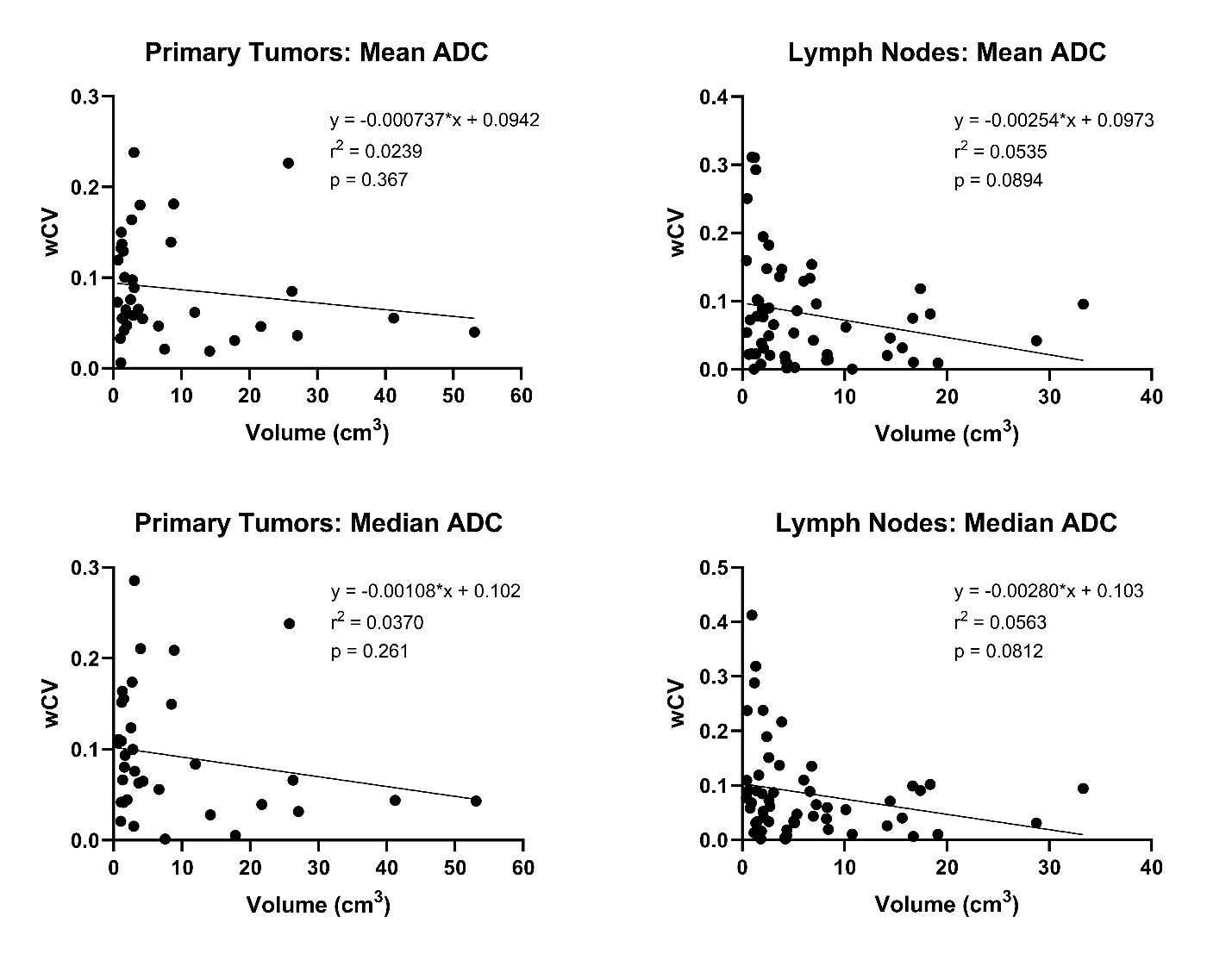

**Figure S2:** wCV vs. volume comparisons for both primary tumors and lymph nodes. wCV values using both mean ADC and median ADC were analyzed. The linear regression equation, r^2^ value, and p-value for the F-test (α=0.05) is shown for each comparison.
